# White Matter Stratification in Depression Predicts Multidimensional Antidepressant Responses

**DOI:** 10.64898/2025.12.08.25341854

**Authors:** Jiaolong Qin, Xinyi Wang, Huangjing Ni, Ye Wu, Haiyan Liu, Lingling Hua, Rui Yan, Hao Tang, Peng Zhao, Zhijian Yao, Qing Lu

## Abstract

**Background:** Major depressive disorder (MDD) is clinically heterogeneous, posing a persistent challenge for personalized treatment. While neuroimaging offers a promising path, existing symptom-based stratification schemes have proven inadequate in predicting antidepressant response. Crucially, studies focusing on white matter (WM) heterogeneity — a potential source of neurobiological subtypes— have failed to address this critical gap. Here, we bridge this divide by investigating WM-based MDD subtypes and their predictive value for treatment outcomes.

**Methods:** We used non-negative matrix factorization biclustering of diffusion MRI data from 311 MDD patients (discovery: n=209; validation: n=102) to identified neuroanatomical subgroups with distinct WM microstructural signatures. Subgroups were characterized via neuroanatomical profiling, clinical phenotyping (symptom domains/treatment responses), and WM-symptom associations. Baseline WM features predicted 4-week treatment outcomes (overall/dimension-specific symptom reduction) across five antidepressant therapies using support vector regression.

**Results:** Three robust MDD subgroups emerged: (1) frontoparietal-corticospinal alterations linked to anxiety/hopelessness; (2) cerebellar-visual circuit disruptions tied to cognitive-psychomotor deficits; (3) fornix-centered abnormalities associated with attenuated symptom severity. Subgroup-specific WM networks predicted treatment outcomes with high cross-cohort consistency (discovery: *r*=0.24–0.58; validation: *r*=0.27–0.67; all *p*<0.05), notably for cognitive symptoms (max *r*=0.59). Importantly, baseline WM patterns—converging on limbic/default mode networks—reflected neuroplasticity reserve, enabling generalizable prediction across mechanistically distinct therapies.

**Conclusions:** Our findings establish WM-derived biotypes as robust, pathophysiologically distinct subtypes of MDD and validate baseline WM topology as a biomarker capable of predicting antidepressant treatment response, potentially by reflecting and individual’s neuroplasticity reserve.

## Introduction

Major depressive disorder (MDD) is a highly heterogeneous condition (American Psychiatric Association, 2013; Fried and Nesse, 2015) with poorly understood pathophysiology (Fried and Nesse, 2015), which sustains a pervasive trial-and-error approach to treatment (Fried, 2017; James et al., 2018). Hence, reliable pretreatment predictors are critically needed to enable personalized therapy, minimize unnecessary medication exposure, and improve resource allocation (Kraus et al., 2019; Thase, 2009). Initial attempts to forecast antidepressant outcomes have primarily depended on symptom-based stratification—utilizing either self-report or clinician-rated scales—to categorize patients and predict treatment response (Arnow et al., 2015; Kato et al., 2020). Although these clinical measures offer practical utility and are widely implemented in routine practice, they provide minimal insight into MDD’s underlying neurobiological mechanisms. This fundamental limitation stems from the non-specific nature of symptoms: identical clinical presentations may arise from disparate biological origins, while shared pathophysiology can manifest as divergent symptom profiles. Consequently, symptom-driven prediction models typically prove inadequate for guiding treatment selection. Illustrating this point, Arnow et al. (Arnow et al., 2015) categorized MDD patients into melancholic, anxious, and atypical subtypes, yet observed significant subgroup overlap and no differential treatment response, ultimately concluding that such classifications possess minimal clinical value for personalized antidepressant selection.

In response to the limitations of symptom-based approaches, research has increasingly turned to neuroimaging as a promising alternative, as it allows for biologically-anchored stratification by mapping symptoms onto discrete neural substrates. The central challenge involves identifying MDD biotypes characterized by distinctive functional or structural brain patterns, thereby facilitating pathophysiology-specific treatments. Crucially, as (Feczko et al., 2019) emphasize, meaningful subtype definitions must correlate with specific clinical or mechanistic outcomes. While pioneer neuroimaging-based endeavors have demonstrated potential for prediction—notably, (Drysdale et al., 2017) delineated four fMRI-based biotypes predictive of transcranial magnetic stimulation response — many existing studies still fail to demonstrate such authentic predictive value, instead predominantly reporting post-hoc correlations (Kennis et al., 2020; Whelan and Garavan, 2014). Furthermore, these problems have been compounded by methodological limitations such as inadequate sample sizes and a lack of external validation (Cohen et al., 2021; Kraus et al., 2019).

Beyond these general methodological issues, this emerging field faces two distinct yet critical limitations that our study aims to address. On one hand, diffusion MRI (dMRI) remains markedly underutilized for MDD subtyping. Despite its direct relevance to neural circuitry, only one study has employed white matter (WM) microstructural abnormalities to characterize heterogeneity (Liang et al., 2019). On the other hand, conventional clustering methodologies dominate current stratification approaches, where derived subtypes’ brain patterns are subsequently obtained through group-level comparative analyses, potentially obscuring intrinsic feature-sample relationships crucial for subtype definition. A noteworthy exception is the fMRI biclustering investigation by (Tokuda et al., 2018), which identified three MDD subtypes with unique functional connectivity patterns. Unlike traditional clustering, biclustering reveals localized associations between specific sample and feature subsets, which may be attenuated in global clustering models using all features.

To overcome these limitations and to translate an improved neurobiological taxonomy into clinically actionable predictors, we examine three key questions: (1) How to stratify MDD subtypes while identifying their unique WM patterns during clustering; (2) How these subtypes differ in their WM profiles and clinical presentations; (3) Whether baseline WM characteristics can predict both overall symptom reduction and domain-specific treatment responses (e.g., anxiety/somatization, cognitive impairment) across diverse therapeutic interventions. The neurobiological rationale for cross-treatment outcome prediction rests on WM pathology serving as a common pathway through which various interventions (e.g., pharmacotherapy, neuromodulation) restore neural circuit function (Long et al., 2025; Mao et al., 2025). Baseline WM architecture reflects neuroplasticity reserve—the inherent capacity for circuit reorganization that underlies treatment response (Castrén and Hen, 2013; Gazerani, 2025; Marzola et al., 2023)—thereby enabling prediction across therapeutic modalities. Our analytical framework addresses these questions through: (1) non-negative matrix factorization (NMF) based biclustering to identify subtype-specific WM-patient relationships; (2) comprehensive characterization of subtypes in discovery and validation cohorts via neuroanatomical profiling and clinical phenotyping; and (3) systematic evaluation of baseline symptom-WM associations, development of baseline WM-based predictive treatment effect models, and assessment of their generalization ability in an external validation dataset.

## Results

### Data-driven MDD subgroup identification

Three clinically distinct and robust subgroups emerged from the discovery cohort analysis, corresponding to the optimal rank *k*=3 (cophenetic correlation value with 0.96; Supplementary Figure S1). Subgroup 1 (n=32) exhibited WM alterations across 5, 730 voxels, subgroup 2 (n=129) involved 5,711 voxels, and subgroup 3 (n=48) comprised 5,566 voxels. Demographic characteristics (e.g., age, sex) showed no significant between-subgroup differences (*p*>0.05, Table 1).

**Table 1.** Clinical characteristics of each subgroup in the discovery dataset.

| Items | Subgroup 1 | Subgroup 2 | Subgroup 3 | *p* value |
| --- | --- | --- | --- | --- |
| Number of subjects | 32 | 121 | 48 | - |
| Age (years) | 32.97±10.77 | 32.80±10.20 | 33.10±7.94 | 0.83<sup>a</sup> |
| Gender (male/female) | 19/13 | 58/71 | 22/26 | 0.33<sup>b</sup> |
| Number of improver/nonimprover | 28/4 | 115/6 | 41/7 | 0.09<sup>b</sup> |
| Total score of 24-item HAMD at baseline | 27.88±7.18 | 29.36±7.83 | 27.17±6.84 | 0.02<sup>a*</sup> |
| Number of recurrent episodes | 2.32±2.67 | 1.80±1.25 | 1.73±1.22 | 0.26<sup>a</sup> |
| The current episode duration (months) | 7.24±4.95 | 5.03±4.15 | 5.16±3.69 | 0.15<sup>a</sup> |
| Total duration of depression (months) | 24.86±23.82 | 28.44±38.73 | 13.10±17.40 | 0.13<sup>a</sup> |
| The effective illness duration (months) | 21.22±21.10 | 9.42±10.11 | 10.08±12.50 | 0.02<sup>a*</sup> |
| Number of improvers in the 5 treatment arms |  |  |  | 0.04<sup>b*</sup> |
| SSRIs monotherapy (n) | 10 | 49 | 21 | - |
| SNRIs monotherapy (n) | 1 | 11 | 2 | - |
| rTMS with SSRIs antidepressant (n) | 8 | 28 | 2 | - |
| rTMS with SNRIs antidepressant (n) | 2 | 9 | 9 | - |
| ECT with antidepressant (n) | 7 | 18 | 7 | - |

Values were expressed as mean±standard deviation. HAMD, Hamilton Depression Rating Scale.
<sup>a</sup> *p*-value was obtained from a Kruskal-wallis analysis of variance (ANOVA).
<sup>b</sup> *p*-value was obtained from Person's $\chi^2$ -test
\* *p*-value was showed a statistical significance.
SSRI = Selective Serotonin Reuptake Inhibitor, SNRI = Serotonin-Norepinephrine Reuptake
Inhibitor, rTMS = Repetitive Transcranial Magnetic Stimulation, ECT = Electroconvulsive Therapy.

In the independent validation cohort, the subgroup stratification demonstrated high robustness, as evidenced by a cophenetic correlation of 0.96. This high value, with <5% variation in individual assignments across 100 NMF iterations, indicates that the derived taxonomy yields consistent and stable clusters. Subgroup distribution mirrored patterns from the discovery phase: Subgroup 1 (n=19), Subgroup 2 (n=64), and Subgroup 3 (n=19), with preserved subgroup-specific WM spatial signatures.

Their demographic characteristics are presented in Table 2. Notably, Subgroup 3 consistently exhibited the lowest mean 24-Hamilton Depression Rating Scale (HAMD) total scores across discovery and validation cohorts, potentially indicating attenuated symptom severity in this neuroanatomical subtype.

**Table 2.** Clinical characteristics of each subgroup in the external independent dataset.

| Items | Subgroup 1 | Subgroup 2 | Subgroup 3 | *p* value |
| --- | --- | --- | --- | --- |
| Number of subjects | 19 | 64 | 19 | - |
| Age (years) | 35.11±12.35 | 32.17±13.09 | 32.05±9.63 | 0.49<sup>a</sup> |
| Gender (male/female) | 6/13 | 26/38 | 9/10 | 0.61<sup>b</sup> |
| Number of improver/nonimprover | 17/2 | 63/1 | 18/1 | 0.20<sup>b</sup> |
| Total score of 24-item HAMD at baseline | 28.64±8.28 | 30.44±9.66 | 27.89±9.34 | 0.72<sup>a</sup> |
| Number of recurrent episodes | 1.82±0.95 | 1.95±1.15 | 1.50±0.79 | 0.30<sup>a</sup> |
| The current episode duration (months) | 3.61±3.04 | 14.27±17.74 | 11.90±14.52 | 0.10<sup>a</sup> |
| Total duration of depression (months) | 50.16±87.19 | 52.38±64.84 | 21.56±19.23 | 0.20<sup>a</sup> |
| The effective illness duration (months) | 6.81±6.84 | 25.69±23.75 | 34.10±11.02 | 0.14<sup>a</sup> |
| Number of improvers in the 5 treatment arms |  |  |  | 0.53<sup>b</sup> |
| SSRIs monotherapy (n) | 1 | 9 | 2 | - |
| SNRIs monotherapy (n) | 1 | 4 | 4 | - |
| rTMS with SSRIs antidepressant (n) | 5 | 11 | 3 | - |
| rTMS with SNRIs antidepressant (n) | 3 | 12 | 4 | - |
| ECT with antidepressant (n) | 2 | 16 | 3 | - |

Values were expressed as mean±standard deviation.
<sup>a</sup> *p*-value was obtained from a Kruskal-wallis analysis of variance (ANOVA).
<sup>b</sup> *p*-value was obtained from Person's $\chi^2$ -test
\* *p*-value was showed a statistical significance.

### Multidimensional subgroup characterization

#### Neuroanatomical characterization of MDD subgroups

The excellent clustering stability facilitates precise identification of subgroup-specific WM signatures (Figure 1). Anatomical location assignment using Natbrain and LNAO_SWM79 atlas (Catani and Thiebaut De Schotten, 2008; Guevara et al., 2017) revealed distinct microstructural profiles (detailed anatomical distribution is provided in Supplementary Table S2):

Subgroup 1: Predominantly superficial fibers concentrated in frontopariental regions (pars opercularis, superior/precentral gyri) with long-association involvement in corticospinal tracts, corpus callosum, and right cingulum/ inferior longitudinal fasciculus (ILF).

Subgroup 2: Cerebellar-focused pathology (cortico-ponto-cerebellar tracts, cerebellar peduncles) combined with distributed long-association alterations (bilateral cingulum/ILF) and diffuse superficial fibers across multiple lobes.

Subgroup 3: Fornix-centered abnormalities with complementary long-association changes (corpus callosum, cingulum) and superficial fibers in parieto-frontotemporal regions.

The analyses of mean fractional anisotropy (FA) value within the subgroup-specific WM signatures demonstrated consistent cross-cohort differences (Figure 2h and 3h). Subgroups 1 and 3 showed elevated FA versus healthy controls (HC) (*p*≤0.001), while Subgroup 2 exhibited reduced FA (*p*<0.004).

#### Clinical phenotypic profiles

##### (1) Clinical symptomatology across MDD subgroups

Analyses of 24-HAMD profiles revealed distinct symptom trajectories across subgroups, and there was a certain degree of agreement in reproducibility between the discovery and validation cohorts (Figure 2a-g, 3a-g). In the discovery cohort, significant between-subgroup differences emerged for 24-HAMD scores (*p*=0.02) and three symptom dimensions: psychomotor retardation (p=0.022), sleep disturbance (*p*=0.002), and hopelessness (*p*=0.043). Radar plot visualization showed that Subgroup 1 was characterized by elevated anxiety/somatization, cognitive impairment, and hopelessness; Subgroup 2, by pronounced cognitive impairment, retardation, and hopelessness; and Subgroup 3, by selective anxiety/somatization elevation.

Validation analyses confirmed core phenotype stability: Retardation (*p*=0.046) and hopelessness (*p*=0.015) remained significantly differentiated. Subgroup-specific severity patterns persisted, with Subgroup 1 remaining characterized by anxiety/somatization and hopelessness; Subgroup 2 by cognitive impairment, retardation, and hopelessness; and Subgroup 3 by anxiety/somatization.

Notably, sleep disturbance differentiation did not replicate in validation, suggesting state-dependent variability. The cross-cohort correlation for symptom profiles is *r* = 0.958 (*p* < 1.951e-08), confirming subgroup consistency.

##### (2) Differential treatment responses across MDD subgroups

Analysis of treatment outcomes revealed distinct response patterns among the three MDD subgroups in both discovery and validation cohorts. In the discovery cohort (Table 1), response rates showed a trend-level difference (*p*=0.09), with Subgroup 2 demonstrating the highest proportion of improvers (115/121, 95.0%) compared to Subgroup 1 (28/32, 87.5%) and Subgroup 3 (41/48, 85.4%). Treatment-stratified analysis revealed significant variation in response patterns (*p*=0.04). Selective Serotonin Reuptake Inhibitor (SSRI) monotherapy responses were most frequent in Subgroup 2 (49 cases). Serotonin-Norepinephrine Reuptake Inhibitor (SNRI) monotherapy showed limited efficacy across all subgroups. Repetitive Transcranial Magnetic Stimulation (rTMS) augmentation demonstrated subgroup-specific effects, particularly when combined with SSRIs in Subgroup 2 (28 cases). Electroconvulsive Therapy (ECT) showed relatively uniform responses across subgroups. Baseline 24-HAMD scores differed significantly between subgroups (*p*=0.02), suggesting varying baseline severity levels. While recurrent episodes and illness duration measures showed no significant differences, effective illness duration was significantly different among subgroups (*p*=0.02).

In the validation cohort (Table 2), response rates were consistently high across all subgroups (89.5 - 98.4%) without significant differences (*p*=0.20). None of the baseline clinical characteristics showed significant inter-subgroup differences in this cohort. Treatment-specific responses were more uniform in validation, with no significant subgroup differences (*p*=0.53), though similar numerical patterns emerged: SSRI monotherapy remained most effective in Subgroup 2 (9 cases); rTMS combinations showed broader efficacy; ECT maintained relatively consistent response rates.

### Treatment outcome prediction

#### Distinct Symptom-WM associative networks across MDD subgroups

The 10 sparse canonical correlation analysis (sCCA) results robustly identified distinct symptom correlates for each subgroup’s WM signature. Subgroup 1 specifically correlated with anxiety/somatization, Subgroup 2 with cognitive impairment and retardation, and Subgroup 3 with all the above three symptom domains (The detailed association results are presented in Supplementary Table S1). Based on these symptom-related WM signatures, affected WM networks were constructed for each subgroup. Figure 4 illustrates the affected WM connectivity patterns observed in each MDD subgroup. Notably, the three subgroups exhibited distinct distributions of altered WM networks: Subgroup 1 primarily demonstrated disruptions in default mode network B, subcortical network, dorsal attention A, limbic networks A/B, salience/ventral attention A, somato-motor A, and central visual network; Subgroup 2 showed predominant alterations in the cerebellar network, default mode networks B/C, limbic A, salience/ventral attention A, dorsal attention A/B, and both peripheral and central visual networks; Subgroup 3 was characterized by WM connectivity changes predominantly involving default mode network B, limbic A, salience/ventral attention A, somato-motor A/B, and visual networks (peripheral and central).

#### Predictive model performance

In the discovery dataset, degree centrality measures derived from affected WM network demonstrated significant predictive capacity for treatment outcomes across all the three subgroups (Figure 5). Specifically, these network-based features successfully forecasted percentage reductions in: 24-HAMD total scores (all subgroups, *r* = 0.28 - 0.46, *p* < 0.05); anxiety/somatization (all subgroups, *r* = 0.38 - 0.58, *p* < 0.007); cognitive impairment (all subgroups, *r* = 0.24 - 0.55, *p* < 0.03); retardation (all subgroups, *r* = 0.33 - 0.52, *p* < 0.009); and hopelessness (all subgroups, *r* = 0.23 - 0.43, *p* < 0.05); as well as sleep disturbance (subgroup 2 and 3, *r* = 0.27 - 0.43, *p* < 0.009).

External validation in an independent cohort (Figure 6) affirmed the generalizability of these findings. Specifically, successful predictions were maintained for percentage reductions in: 24-HAMD total score (all subgroups, *r* = 0.28 - 0.60, *p* < 0.042), cognitive impairment (all subgroups, *r* = 0.29 - 0.59, *p* < 0.03), anxiety/somatization (Subgroup 1 and 2, *r* = 0.27 - 0.53, *p* < 0.04), retardation (Subgroup 2 and 3, *r* = 0.30 - 0.67, *p* < 0.03), and sleep disturbance (Subgroup 2, *r* = 0.27, *p* = 0.043), as well as feeling of hopelessness (Subgroup 1, *r* = 0.62, *p* = 0.021).

## Discussion

This study advances the neurobiological parsing of MDD heterogeneity by identifying three distinct subgroups with differential WM and clinical profiles. By identifying three robust MDD biotypes through NMF biclustering (cophenetic value = 0.96), we demonstrate that pre-treatment WM topology contains predictive information that is generalizable across independent cohorts and, crucially, across mechanistically distinct treatments. The consistent predictive accuracy (discovery: r = 0.24–0.58; validation: r = 0.27–0.67; all p < 0.05) suggests that these baseline WM patterns reflect a stable neurobiological trait—most plausibly, an individual’s neuroplasticity reserve—which underlies and constrains the brain’s capacity for functional recovery regardless of the specific therapeutic intervention.

### Neuroanatomical subgroups and pathophysiological implications

Our identification of three neuroanatomical MDD subgroups, characterized by unique WM disruption patterns, provides novel insights into the heterogeneous pathophysiology of depression. Subgroup 1 exhibited frontoparietal-corticospinal alterations, predominantly involving superficial WM fibers—a finding contrasting with traditional deep WM-focused MDD studies. These disruptions align with this subgroup’s prominent anxiety and hopelessness, implicating dysregulated default mode and salience networks in emotional dysregulation (Menon, 2011). Subgroup 2 demonstrated cerebellar-visual circuit abnormalities, including the ILF, suggesting disrupted cerebro-cerebellar loops may underlie its cognitive-psychomotor deficits (Schmahmann et al., 2019). Subgroup 3 showed fornix-centered alterations, a structure integral to memory and reward processing (Benear et al., 2020; Godsil et al., 2013), potentially reflecting impaired hippocampal-prefrontal connectivity associated with attenuated symptom severity.

Notably, all subgroups shared disruptions in major WM tracts—corpus callosum, cingulum, and ILF—with distinct spatial patterns. The corpus callosum, the largest inter-hemisphere association, may be related to the pathogenesis of depressive symptoms in psychiatric disorders including MDD (Cole et al., 2012). The cingulum forms the outer ring of the limbic WM tracts, which are involved in key brain functions such as emotion, executive function, and episodic memory (Bubb et al., 2018; Pascalau et al., 2018). The ILF connects the occipital cortex to the anterior temporal region. As the WM backbone of the ventral visual pathway, it is considered crucial for maintaining a variety of cognitive and emotional processes in the visual modality, particularly in object and face recognition, visual emotion recognition, language, and semantics (Herbet et al., 2018; Zemmoura et al., 2021). A study by (Haghshomar et al., 2018) summarized the associations between the ILF and the symptoms of Parkinson’s disease, and suggested that this bundle is involved in the recognition of negative facial emotions in its symptoms related to mood disorders. Prior studies have reported the disruptions to these three WM bundles in MDD (Liao et al., 2013; Luttenbacher et al., 2022; Pasternak et al., 2018). Our results are consistent with previous findings, but we further reveal that damage to these bundles varies in specific locations and that FA value changes differ across different subgroups. Specifically, the mean FA value is decreased in Subgroup 2, while it is increased in Subgroup 1 and 3. These findings suggest that distinct sub-bundles exist within these bundles and play different roles in the pathological mechanisms of MDD.

### WM stratification in MDD predicts symptom-specific treatment outcomes

Our study demonstrates that WM-based stratification directly predicts overall antidepressant outcomes and domain-specific improvements (especially in cognitive symptom) across subgroups. This neuroanatomical approach moves beyond the limitations of conventional, symptom-based subtyping, which often fails to inform treatment selection due to its inability to capture underlying pathophysiology (Arnow et al., 2015; Kato et al., 2020; Kautzky et al., 2021). In contrast, our WM-derived subgroups exhibit distinct neurostructural homogeneity, enabling a direct link between neuroanatomical pathology, clinical symptomatology, and treatment outcomes. By establishing this brain-symptom-outcome relationship, our work provides a pathophysiology-grounded framework for predicting treatment effects, thereby delivering actionable biomarkers to guide personalized therapy selection.

Our findings demonstrate symptom-specific neurobiological substrates underlying antidepressant treatment prediction across three depression subgroups. In Subgroup 2, cerebellar-visual circuit impairment predicted outcomes in five dimensions (overall efficacy, anxiety-somatization, cognitive impairment, psychomotor retardation, sleep disturbance), excluding hopelessness. This finding provides further support for the concept that cerebellar-visual abnormalities disrupt emotional-cognitive integration via two pathways: (1) amplifying negative visual biases via dysregulated limbic modulation (Ciapponi et al., 2023), and (2) impairing attention/visuospatial processing through compromised cerebellar-prefrontal interactions (Yildiz et al., 2010). Subgroup 1 exhibited frontoparietal-corticospinal alterations uniquely predicting hopelessness alongside anxiety-somatization and cognitive deficits. These associations may stem from disrupted executive control (frontoparietal network dysfunction) and failed emotional-somatic integration, where aberrant prefrontal-limbic connectivity biases reward processing and interoception, sustaining negative future expectations (Kaiser et al., 2015). In contrast, Subgroup 3 showed multi-tract WM pathology involving fornix, corpus callosum, cingulum, and ILF, specifically linked to severe cognitive impairment and psychomotor retardation. These structural disruptions form a disconnection syndrome that manifests as cognitive deficits through impaired information integration and psychomotor symptoms via disrupted motivation-motor coordination.

The baseline imaging data of the three subgroups can predict the treatment outcomes at 4 weeks in a mixed cohort of MDD patients receiving at least one of five antidepressant therapies. Prior studies reported that pretreatment baseline neuroimaging features could predict treatment response across diverse interventions (e.g., pharmacotherapy, neuromodulation therapy, and psychotherapy) (Long et al., 2025; Mao et al., 2025). Nevertheless, whether the imaging features predict outcomes generally or differ depending on specific treatments remains unclear. Our results support the generalizability of baseline imaging biomarkers. This cross-treatment validity may stem from shared neural mechanisms in MDD, as predictive features consistently involved limbic and default mode network, suggesting different therapies modulate common brain circuits. Ultimately, our findings imply that the pretreatment degree metrics of symptom-related network reflect robust network-level disturbances in MDD, capturing core pathophysiological states that influence treatment response beyond specific therapeutic modalities. These pretreatment features could serve as an initial screening tool to identify patients with a high risk of treatment resistance, avoiding unnecessary trial-and-error in clinical practice.

### Data-driven identification of MDD subgroups through NMF-biclustering

We employed an NMF-based biclustering approach to identify localized WM alterations in MDD, yielding three robust WM patterns. These data-driven patterns demonstrated: (1) significant regional FA differences compared to HC; (2) strong associations with core depressive symptoms (*r* = 0.52 ∼ 0.92; supplementary Table S1); (3) predictive value for treatment outcomes.

This biclustering approach offers key advantages over conventional monoclustering method in the task of subtyping MDD: (1) captures localized WM alterations and preserves spatial heterogeneity through simultaneous sample-feature clustering; (2) identifies subgroup-specific features without requiring HC comparisons, avoiding the “averaging problem” inherent in classical subgroup stratification by the monoclustering method. This methodological advance enables more precise mapping of the neurobiological heterogeneity in MDD by identifying localized co-variation patterns that would be obscured in conventional case-control analyses.

### Limitations

The limitations of our study include its reliance solely on WM features, which overlooks potential interactions with other modalities like cortical thickness or functional connectivity that could refine subtype characterization (Drysdale et al., 2017). Additionally, the coarse resolution of current WM atlases limits precise localization of alterations within fiber bundles, necessitating finer parcellation schemes. Future studies should employ multimodal imaging, larger multi-site cohorts, and standardized protocols to dissect confounding factors (e.g., scanner variability, sociogeographic influences) and validate the predictive models’ robustness.

## Conclusion

Our study demonstrates that data-driven analysis of WM architecture robustly parses the heterogeneity of MDD into neurobiologically distinct biotypes. These subtypes are characterized by specific neuroanatomical signatures and clinical symptom profiles. The predictive power of baseline WM topology for treatment outcomes underscores its potential as an actionable biomarker, and their WM patterns may serve as a proxy for an individual’s neuroplasticity reserve, offering a mechanistic explanation for treatment efficacy.

## Materials and methods

### Participants

From March 2012 to July 2018, 311 MDD patients were recruited from Nanjing Brain Hospital (n=209, discovery dataset) and Nanjing Drum Tower Hospital (n=102, validation dataset). All patients met DSM-IV criteria (Association, 1994) as confirmed by the Chinese version of MINI, and their depressive symptoms were assessed using 24-HAMD (Hamilton, 1960). Medication-treated patients completed a 2-week washout period prior to baseline. Two trained psychiatrists conducted the 24-HAMD assessments at baseline and after 4 weeks of treatment. We also enrolled 100 age-, gender-, and handedness-matched HC from local communities. The demographic and clinical characteristics are presented in Table 3.

**Table 3.** Demographic characteristic and clinical information of depressions and healthy controls in the current study.

| Items | Discovery dataset<br>(Nanjing Brain Hospital) | External independent dataset<br>(Nanjing Drum Tower Hospital) | HC dataset<br>(Nanjing Brain Hospital) | *p* value |
| --- | --- | --- | --- | --- |
| Number of subjects | 209 | 102 | 100 | - |
| Age (years) | 32.89±9.77 | 32.70±12.33 | 31.80±8.45 | 0.34<sup>a</sup> |
| Gender (male/female) | 99/110 | 41/61 | 44/56 | 0.58<sup>b</sup> |
| Handedness (right/left) | 209/0 | 102/0 | 100/0 | 0.99<sup>b</sup> |
| Total score of 24-item HAMD at baseline | 28.63±7.54 | 29.64±9.33 | - | - |
| Total score of 24-item HAMD after 4-week | 9.26±7.47 | 12.42±7.57 | - | - |
| Number of recurrent episodes | 1.86±1.54 | 1.84±1.06 | - | - |
| The current episode duration (months) | 8.99±15.07 | 11.94±15.84 | - | - |
| Total duration of depression (months) | 34.18±51.45 | 45.87±64.34 | - | - |
| The effective illness duration (months) | 12.72±17.32 | 24.17±20.84 | - | - |
| Treatment arms |  |  |  |  |
| SSRIs monotherapy (%) | 43.88 |  | - | - |
| SNRIs monotherapy (%) | 7.65 |  | - | - |
| rTMS with SSRIs antidepressant (%) | 19.39 |  | - | - |
| rTMS with SNRIs antidepressant (%) | 11.22 |  | - | - |
| ECT with antidepressant (%) | 17.86 |  | - | - |
| Antidepressant mean dosage (mg/day) |  |  |  |  |
| SSRIs monotherapy |  |  |  |  |
| Escitalopram | 16.81±4.34 | 17.89±3.46 | - | - |
| Sertraline | 150±58.63 | 131.82±40.45 | - | - |
| Fluoxetine | 20±0 | 20±0 | - | - |
| Paroxetine | 28.33±7.53 | 35±8.66 | - | - |
| Fluvoxamine | 275±41.83 | 162.5±25 | - | - |
| SNRIs monotherapy |  |  |  |  |
| Venlafaxine | 193.63±42.55 | 205±33.73 | - | - |
| Duloxetine | 90±34.64 | 76±26.08 | - | - |

Values were expressed as mean±standard deviation. HC = Healthy controls.
<sup>a</sup> *p*-value was obtained from an independent two-sample *t*-test between discovery dataset and HC dataset.
<sup>b</sup> *p*-value was obtained from Person's $\chi^2$ -test between discovery dataset and HC dataset.

The treatment strategies, determined by two experienced clinicians based on patients’ history and symptoms, included: (1) SSRIs monotherapy, (2) SNRIs monotherapy, (3) dual antidepressant therapy (SSRI+SNRI), (4) antidepressant combined with ECT therapy, or (5) antidepressant complemented by rTMS.

Exclusion criteria included: (1) substance abuse/dependence or major medical disorders, (2) history of neurological conditions or brain injury, and (3) MRI contraindications. All participants provided written informed consent after receiving detailed study information. The study protocol was approved by the Ethics Review Boards of both Nanjing Brain Hospital and Nanjing Drum Tower Hospital in accordance with the Declaration of Helsinki.

### Imaging acquisitions and preprocessing

All MRI data were acquired using 3.0 T Siemens Verio scanners equipped with 12-channel head coils at both study sites. Consistent imaging protocols were implemented across sites for both dMRI and T1-weighted acquisitions. DMRI parameters included: 30 diffusion directions (b=1,000 s/mm^2^) with one b0 image, TR/TE=6600/93 ms, flip angle = 90^◦^, FOV = 240 × 240 mm^2^, matrix = 128 × 128, 3 mm slice thickness with no gap, and voxel size= 1.9 × 1.9 × 3 mm^3^. T1-weigthed imaging parameters were: TR/TE = 1900/2.48 ms, 1 mm slice thickness, flip angle = 9^◦^, inversion time = 900 ms, matrix = 256 × 256, FOV = 250 × 250 mm^2^, and isotropic 1 mm^3^ voxels.

All images underwent visual inspection for artifacts before standardized preprocessing. For both T1w and dMRI data, we performed: (1) axial alignment; (2) Gibbs ringing correction using local Subvoxel-Shifts (Kellner et al., 2016); and (3) N4ITK intensity inhomogeneity correction (Tustison et al., 2010). DMRI-specific processing included: (1) MP-PCA denoising (Veraart et al., 2016); (2) eddy current correction (FSL’s eddy_correct tool) (Jenkinson et al., 2012); (3) EPI correction; (4) CNN-based brain masking (pnlNipype); (5) T1w-dMRI alignment used FSL (Jenkinson et al., 2012) rigid registration while preserving native diffusion space. (6) Diffusion tensors were computed using the Stejskal-Tanner equation. Eigenvalue decomposition yielded fractional anisotropy (FA) maps.

Whole-brain probabilistic tractography was performed in MRtrix3 using the iFOD2 algorithm, with fiber orientation distributions first estimated using constrained spherical deconvolution. A total of 3 million seeds were employed to initiate streamline propagation, with all other tracking parameters set to their default values (Tournier et al., 2019). Subsequently, the resulting tractograms were processed using the Spherical-deconvolution Informed Filtering of Tractograms (SIFT) method (Smith et al., 2015, 2013) to obtain a refined and biologically meaningful set of 1 million streamlines.

For Tract Based Spatial Statistics (TBSS) analysis in FSL 6.0 (Smith et al., 2006), individual FA maps were nonlinearly registered to the FMRIB58_FA template. A mean FA skeleton was generated, thresholded at FA>0.2 to exclude partial volume effects, representing core WM tracts. Each subject’s FA data were projected onto this skeleton by identifying maximal FA values along directions perpendicular to the skeleton. This standardized approach ensured comparable WM analysis across all participants.

### Data-driven identification of MDD subgroups

#### Biclustering identification via NMF in discovery dataset

Each patient’s FA-TBSS map was flattened into a feature vector, retaining only skeleton voxels. These vectors were then combined to form matrix *X* (109, 848 voxels × 209 subjects). Prior to NMF-based biclustering, we regressed out age and sex covariates from the *X* matrix to control for potential confouding variables. We employed the sparse NMF algorithm from Li (Li and Ngom, 2013) to decompose the TBSS-derived FA matrix *X* ∈ *R^p^*^×^*^n^* (p= voxels, n=subjects). The *X* was factorized into basis matrix *A* ∈ *R^p^*^×^*^k^* (*k* latent components, and its value ranged from 2 to 8 here) and coefficient matrix *Y* ∈ *R^k^*^×^*^n^*, such that *X* ≈ *AY*. Each column of *A* represents a latent WM microstructural pattern formed by voxel combinations, while *Y* quantifies subject-specific expression levels of these patterns. Subject clustering was determined by identifying the dominant component (maximum value) in each column of *Y*, with basis vectors serving as cluster centroids.

To enhance feature interpretability, we reduced feature redundancy through sparsity constraints during decomposition and performed entropy-based feature selection on matrix *A*, retaining the top 20% voxels with highest discriminative power for subsequent biclustering. This dual optimization process simultaneously identifies: (a) cohesive WM patterns through column-wise clustering of *A*, and (b) patient subgroups via row-wise clustering of *Y* (Detailed implementation parameters are provided in Supplementary Methods Section: *identification of biclusters based on NMF method)*.

To determine the optimal rank *k* that yields stable and meaningful clusters, we adopted Brunet’s consensus clustering approach (Brunet et al., 2004). For each rank *k*, we performed 100 iterations of matrix factorization to account for random initialization effects. Each iteration generated a connectivity matrix *C* (n × n), where *C_ij_* =1 if samples *i* and *j* clustered together and 0 otherwise. The consensus matrix was computed by averaging these 100 connectivity matrices, with entries representing the probability of pairwise sample co-clustering. We then performed hierarchical clustering (average linkage) on the consensus matrix and evaluated clustering stability using the cophenetic correlation coefficient, which quantifies the agreement between the consensus matrix distances and hierarchical clustering linkage distances. The optimal *k* was selected as the value maximizing this coefficient (Figure 7).

#### Bicluster assignment in independent validation dataset

To classify new patients from the validation cohort, we projected each individual’s data onto the pre-established basis matrix *A* to compute coefficient matrix *Y*, where maximal values in each column determine cluster membership. This assignment process was repeated 100 times to generate a test consensus matrix, mirroring the protocol from the discovery phase. Hierarchical clustering with average linkage was then applied, followed by computation of the corresponding cophenetic correlation coefficient. This was done to assess the stability of the clustering using the predetermined optimal rank (*k*) when applied to the validation cohort.

### Subgroup characterization analysis

#### WM signature extraction

To identify subgroup-specific WM microstructural signatures, we developed a frequency-based feature selection protocol following NMF biclustering. Given the stochastic nature of biclustering assignments across 100 iterations (performed at the predetermined optimal rank *k*), we quantified voxel-wise feature consistency through the following steps (Figure 8):

1. **Feature occurrence mapping**: Created a frequency matrix *F* (sample pairs × features) by incrementing entries when features co-occurred in biclusters across iterations, normalized to 0 - 1 probability values. Specifically, each row represented a unique sample pair, and each column corresponded to a voxel feature. For each biclustering run, if a feature (column) appeared in the same cluster as a sample pair (row), its entry in *F* was incremented by 1.
2. **Consensus-driven partitioning**: Segmented *F* into *k* submatrices by grouping rows (sample pairs) according to their subgroup labels derived from the hierarchical clustering.
3. **Signature thresholding**: For each subgroup-specific submatrix, retained features (columns) exceeding probability threshold (*p* > 0.9) were defined as characteristic WM signatures, ensuring > 90% consistency across biclustering realizations.

This approach transformed each run of biclustering solutions into spatially stable WM profiles, mitigating initialization-dependent variability inherent to NMF decomposition.

#### FA-based case-control comparisons

To statistically validate the identified WM patterns, we conducted comparative analysis of mean FA values between each MDD subgroup and HCs within the signature WM profiles. These comparisons were performed independently in both discovery and validation cohorts using Wilcoxon rank-sum tests. Prior to analysis, we accounted for potential confounding effects by regressing out age and sex covariates.

#### Clinical phenotype differentiation

We conducted comprehensive clinical characterization of the MDD subgroups across discovery and validation datasets. First, we evaluated differences in depression severity (total 24-HAMD score) and symptom domain profiles (anxiety/somatization, cognitive impairment, psychomotor retardation, sleep disturbance, and hopelessness factor scores) (See Supplementary Materials for detailed definitions of symptom factors) using Kruskal-Wallis tests. Second, we examined illness course characteristics including recurrence frequency, current episode duration, total illness duration, and effective illness duration. Significant findings underwent post-hoc analysis using Wilcoxon rank-sum tests in both datasets. Before conducting the above comparisons, we regressed out age and sex effects. Third, we assessed treatment response heterogeneity by comparing responder rates (≥ 20% 24-HAMD reduction after 4 weeks) (Kennedy, 2022) across subgroups for five treatment strategies, analyzed by chi-square tests.

### Predicting antidepressant treatment outcomes across MDD subgroups

#### Symptom-WM associations in discovery cohort

We employed sCCA to investigate associations between the 7 clinical symptom profiles (the above-mentioned 5 symptom domain profiles, plus weight and diurnal variations) and WM microstructural patterns within each MDD subgroup. The analytical pipeline comprised four stages:

1. **Dimensionality reduction**: Given the high feature-to-sample ratio (> 5,000 voxels vs. Subgroup sizes), we implemented region-of-interest (ROI) clustering to address dimensionality. WM patterns were parcellated into spatially contiguous clusters (10 - 20 voxels) (Zalesky et al., 2010), with FA values averaged within each cluster. This ROI-based feature extraction was repeated to 10 times per subgroup to account for clustering stochasticity.
2. **sCCA implementation**: Reduced-dimension data were analyzed via sCCA with cross-validation (CV) to identify maximally correlated linear combinations between clinical factors and WM features, generating canonical variate pairs.
3. **Statistical validation**: Permutation testing (1, 000 iterations) assessed the significance of canonical correlations against null distributions.
4. **Feature stability assessment**: Bootstrapping (1, 000 resamples) identified consistently contributing clinical and neuroimaging features across sCCA solutions.

To assess the robustness of our findings, the established sCCA workflow (Xia et al., 2018) was iterated over 10 different ROI parcellations per subgroup. We examined the stability of clinical loadings and model performance in held-out data. The technical details are provided in Supplementary Methods.

Following sCCA analysis, we identified WM ROI (WM_cca_) associated with 7 clinical symptom factors in each subgroup, retaining ROI appearing in ≥ 2/10 iterations. Fiber tracts passing through these WM_cca_ regions were reconstructed into subgroup-specific WM networks (288 nodes, nodal definitions in Supplementary Excel 1 and are based on the three atlas: Yeo 17 - network, SUIT cerebellum, Tian’s subcortex (Diedrichsen et al., 2009; Schaefer et al., 2018; Tian et al., 2020), representing symptom-related neurocircuitry.

#### Subgroup-specific treatment outcome prediction

The nodes in the symptom-related network were categorized into 19 functional subsystems (e.g., Yeo 17 functional networks, cerebellum network, and subcortical network). Degree centrality values for each subsystem (Calculation details in Supplementary Methods) served as predictive features in support vector regression (SVR) models to forecast six treatment outcomes (e.g., percentage reductions in 24-HAMD total score and five core symptom domain profiles). All features were separately adjusted for age/sex covariates in both datasets prior to the prediction.

Model development employed epsilon-insensitive SVR with sigmoid kernel (Chang and Lin, 2011). Hyperparameter optimization via grid search explored: γ(default=1), coef0 (0.1 - 2, step = 0.01), ε (0.01 - 2, step = 0.01). Prediction performance was evaluated through nested leave-one-out CV (LOOCV), with grid search external to CV folds to prevent data leakage. Final model performance was quantified via Pearson correlation between predicted and observed values across all LOOCV iterations.

#### Model validation in independent cohort

The 18 subgroup-specific SVR models (3 subgroups × 6 treatment outcomes) trained in the discovery cohort were rigorously validated using the independent dataset. Fixed hyperparameters, optimized in the discovery phase, were retained during training of the SVR models. The external independent dataset underwent an identical feature preprocessing pipeline (including age/sex regression and degree centrality calculation) as applied to the discovery cohort. Model performance was quantified using the correlation between predicted and observed values for each model. This validation framework ensured generalizability while preventing overfitting through a complete separation between training and validation datasets.

## Data Availability

All data produced in the present study are available upon reasonable request to the authors.

## Acknowledgment

Sincere appreciation is extended to the patients and control subjects for their valuable participation. We sincerely thank to Professor Chen Gong for his valuable suggestions of NMF-clustering method.

## CRediT Author Statement

Jiaolong Qin: Conceptualization, Methodology, Software, Formal analysis, Validation, Writing - Original Draft, Writing - Review & Editing

Xinyi Wang, Huangjing Ni, Ye Wu: Formal analysis, Data Curation

Haiyan Liu, Lingling Hua, Rui Yan, Hao Tang, Peng Zhao: Conceptualization, Investigation, Resources, Data Curation

Zhijian Yao: Conceptualization, Resources, Supervision

Qing Lu: Conceptualization, Methodology, Supervision, Writing - Review & Editing

## Compliance with ethical standards

### Ethical approval

All procedures performed in studies involving human participants were in accordance with the ethical standards of the institutional and/or national research committee and with the 1964 Helsinki declaration and its later amendments or comparable ethical standards.

Figure 1. The results of WM signature extraction of the three subgroups. a), b) and c) parts presented the coronal brain plane of subgroup 1, 2 and 3, respectively.

Figure 2. The results of clinical symptomatology and mean FA values across MDD subgroups in the discovery dataset. a) Five symptom dimensions were visualized using radar chart. b-f) Between-group differences in scores for five symptom—anxiety/somatization, cognitive impairment, retardation, sleep disturbance and feeling of hopelessness—across the three subgroups were shown in panels b to f, respectively. h) Comparison of the mean FA values of the three WM signature patterns between each subgroup and healthy controls.

Figure 3. The results of clinical symptomatology and mean FA values across MDD subgroups in the external validation dataset. a) Five symptom dimensions were visualized using radar chart. b-f) Between-group differences in scores for five symptom—anxiety/somatization, cognitive impairment, retardation, sleep disturbance and feeling of hopelessness—across the three subgroups were shown in panels b to f, respectively. h) Comparison of the mean FA values of the three WM signature patterns between each subgroup and healthy controls.

Figure 4. Brain WM networks associated with major clinical symptoms at the average group-level. The thickness of each connection reflected the proportion of edge presence across subgroups. TempPar - temporal parietal, DefaultC - default C, DefaultB - default B, DefaultA - default A, ContC - control C, ContB - control B, ContA - control A, LimbicA - limbic A, LimbicB - limbic B, SalVentAttnB - salience/ventral attention B, SalVentAttnA - salience/ventral attention A, DorsAttnB - dorsal attention B, DorsAttnA - dorsal attention A, SomMotB - somatomotor B, SomMotA - somatomotor A, VisPeri - peripheral visual, VisCent - central visual, Subcor - subcortical network.

Figure 5. Prediction of antidepressant treatment outcomes using degree centrality of affected WM network in the discovery dataset. Columns represented six antidepressant treatment outcome measures: percentage reduction in 24-HAMD total score and five core symptom domains—anxiety/somatization, cognitive impairment, retardation, sleep disturbance, and feeling of hopelessness. Rows indicated subgroups (*i.e.*, 1, 2 and 3). * *p*≤0.05, ** *p*≤0.01, *** p≤0.001

Figure 6. Prediction of antidepressant treatment outcomes using degree centrality of affected WM network in the external validation dataset. Columns represented six antidepressant treatment outcome measures: percentage reduction in 24-HAMD total score and five core symptom domains—anxiety/somatization, cognitive impairment, retardation, sleep disturbance, and feeling of hopelessness. Rows indicated subgroups (*i.e.*, 1, 2 and 3). N/A indicated the absence of a valid SVR model applicable to the external dataset. * *p*≤0.05, ** *p*≤0.01, *** p≤0.001

Figure 7. The pipeline of stratifying subgroup in the discovery dataset. First, the TBSS method was used to extract FA values within the WM skeleton. NMF bi-clustering was then performed across a range of predefined ranks *k* (e.g., 2 ∼ 8) to generate corresponding clustering results (e.g., 2 ∼ 8 subgroups). Finally, the cophenetic correlation coefficient was employed to determine the optimal number of subgroups.

Figure 8. The flowchart of WM signature extraction. a) Feature occurrence mapping was illustrated how to generate the frequency matrix F. b) The split of matrix F involved two parts: consensus-driven partitioning and signature thresholding.

## References

American Psychiatric Association, DSM-5 Task Force. 2013. Diagnostic and statistical manual of mental disorders: DSM-5TM, 5th ed. Diagnostic and statistical manual of mental disorders: DSM-5TM, 5th edAmerican Psychiatric Publishing, Inc. DOI: 10.1176/appi.books.9780890425596

Arnow BA, Blasey C, Williams LM, Palmer DM, Rekshan W, Schatzberg AF, Etkin A, Kulkarni J, Luther JF, Rush AJ. 2015. Depression Subtypes in Predicting Antidepressant Response: A Report From the iSPOT-D Trial. American Journal of Psychiatry 172:743–750. DOI: 10.1176/appi.ajp.2015.14020181

Association AP. 1994. Diagnostic Criteria from DSM-IV. American Psychiatric Association.

Benear SL, Ngo CT, Olson IR. 2020. Dissecting the Fornix in Basic Memory Processes and Neuropsychiatric Disease: A Review. Brain Connectivity 10:331–354. DOI: 10.1089/brain.2020.0749

Brunet J-P, Tamayo P, Golub TR, Mesirov JP. 2004. Metagenes and molecular pattern discovery using matrix factorization. Proceedings of the National Academy of Sciences 101:4164–4169. DOI: 10.1073/pnas.0308531101

Bubb EJ, Metzler-Baddeley C, Aggleton JP. 2018. The cingulum bundle: Anatomy, function, and dysfunction. Neuroscience & Biobehavioral Reviews 92:104–127. DOI: 10.1016/j.neubiorev.2018.05.008

Castrén E, Hen R. 2013. Neuronal plasticity and antidepressant actions. Trends in Neurosciences 36:259–267. DOI: 10.1016/j.tins.2012.12.010

Catani M, Thiebaut De Schotten M. 2008. A diffusion tensor imaging tractography atlas for virtual in vivo dissections. Cortex 44:1105–1132. DOI: 10.1016/j.cortex.2008.05.004

Chang C-C, Lin C-J. 2011. LIBSVM: A library for support vector machines. ACM Trans. Intell. Syst. Technol. 2:27:1–27:27. DOI: 10.1145/1961189.1961199

Ciapponi C, Li Y, Osorio Becerra DA, Rodarie D, Casellato C, Mapelli L, D’Angelo E. 2023. Variations on the theme: focus on cerebellum and emotional processing. Frontiers in Systems Neuroscience 17. DOI: 10.3389/fnsys.2023.1185752

Cohen SE, Zantvoord JB, Wezenberg BN, Bockting CLH, van Wingen GA. 2021. Magnetic resonance imaging for individual prediction of treatment response in major depressive disorder: a systematic review and meta-analysis. Translational Psychiatry 11:1–10. DOI: 10.1038/s41398-021-01286-x

Cole J, Chaddock CA, Farmer AE, Aitchison KJ, Simmons A, McGuffin P, Fu CHY. 2012. White matter abnormalities and illness severity in major depressive disorder. The British Journal of Psychiatry 201:33–39. DOI: 10.1192/bjp.bp.111.100594

Diedrichsen J, Balsters JH, Flavell J, Cussans E, Ramnani N. 2009. A probabilistic MR atlas of the human cerebellum. NeuroImage 46:39–46. DOI: 10.1016/j.neuroimage.2009.01.045

Drysdale AT, Grosenick L, Downar J, Dunlop K, Mansouri F, Meng Y, Fetcho RN, Zebley B, Oathes DJ, Etkin A, Schatzberg AF, Sudheimer K, Keller J, Mayberg HS, Gunning FM, Alexopoulos GS, Fox MD, Pascual-Leone A, Voss HU, Casey BJ, Dubin MJ, Liston C. 2017. Resting-state connectivity biomarkers define neurophysiological subtypes of depression. Nature Medicine 23:28–38. DOI: 10.1038/nm.4246

Feczko E, Miranda-Dominguez O, Marr M, Graham AM, Nigg JT, Fair DA. 2019. The Heterogeneity Problem: Approaches to Identify Psychiatric Subtypes. Trends in Cognitive Sciences 23:584–601. DOI: 10.1016/j.tics.2019.03.009

Fried E. 2017. Moving forward: how depression heterogeneity hinders progress in treatment and research. Expert Review of Neurotherapeutics 17:423–425. DOI: 10.1080/14737175.2017.1307737, PMID: 28293960

Fried EI, Nesse RM. 2015. Depression is not a consistent syndrome: An investigation of unique symptom patterns in the STAR*D study. Journal of Affective Disorders 172:96–102. DOI: 10.1016/j.jad.2014.10.010

Gazerani P. 2025. The neuroplastic brain: current breakthroughs and emerging frontiers. Brain Research 1858:149643. DOI: 10.1016/j.brainres.2025.149643

Godsil BP, Kiss JP, Spedding M, Jay TM. 2013. The hippocampal–prefrontal pathway: The weak link in psychiatric disorders? European Neuropsychopharmacology 23:1165–1181. DOI: 10.1016/j.euroneuro.2012.10.018

Guevara M, Román C, Houenou J, Duclap D, Poupon C, Mangin JF, Guevara P. 2017. Reproducibility of superficial white matter tracts using diffusion-weighted imaging tractography. NeuroImage 147:703–725. DOI: 10.1016/j.neuroimage.2016.11.066

Haghshomar M, Dolatshahi M, Ghazi Sherbaf F, Sanjari Moghaddam H, Shirin Shandiz M, Aarabi MH. 2018. Disruption of Inferior Longitudinal Fasciculus Microstructure in Parkinson’s Disease: A Systematic Review of Diffusion Tensor Imaging Studies. Frontiers in Neurology 9. DOI: 10.3389/fneur.2018.00598

Hamilton M. 1960. A Rating Scale for Depression. Journal of Neurology, Neurosurgery & Psychiatry 23:56–62. DOI: 10.1136/jnnp.23.1.56, PMID: 14399272

Herbet G, Zemmoura I, Duffau H. 2018. Functional Anatomy of the Inferior Longitudinal Fasciculus: From Historical Reports to Current Hypotheses. Frontiers in Neuroanatomy 12. DOI: 10.3389/fnana.2018.00077

James SL, Abate D, Abate KH, et al. 2018. Global, regional, and national incidence, prevalence, and years lived with disability for 354 diseases and injuries for 195 countries and territories, 1990–2017: a systematic analysis for the Global Burden of Disease Study 2017. The Lancet 392:1789–1858. DOI: 10.1016/S0140-6736(18)32279-7, PMID: 30496104

Jenkinson M, Beckmann CF, Behrens TEJ, Woolrich MW, Smith SM. 2012. FSL. NeuroImage 62:782–790. DOI: 10.1016/j.neuroimage.2011.09.015

Kaiser RH, Andrews-Hanna JR, Wager TD, Pizzagalli DA. 2015. Large-Scale Network Dysfunction in Major Depressive Disorder: A Meta-analysis of Resting-State Functional Connectivity. JAMA Psychiatry 72:603–611. DOI: 10.1001/jamapsychiatry.2015.0071

Kato M, Asami Y, Wajsbrot DB, Wang X, Boucher M, Prieto R, Pappadopulos E. 2020. Clustering patients by depression symptoms to predict venlafaxine ER antidepressant efficacy: Individual patient data analysis. Journal of Psychiatric Research 129:160–167. DOI: 10.1016/j.jpsychires.2020.06.011

Kautzky A, Möller H-J, Dold M, Bartova L, Seemüller F, Laux G, Riedel M, Gaebel W, Kasper S. 2021. Combining machine learning algorithms for prediction of antidepressant treatment response. Acta Psychiatrica Scandinavica 143:36–49. DOI: 10.1111/acps.13250

Kellner E, Dhital B, Kiselev VG, Reisert M. 2016. Gibbs-ringing artifact removal based on local subvoxel-shifts. Magnetic Resonance in Medicine 76:1574–1581. DOI: 10.1002/mrm.26054

Kennedy SH. 2022. Beyond Response: Aiming for Quality Remission in Depression. Advances in Therapy 39:20–28. DOI: 10.1007/s12325-021-02030-z

Kennis M, Gerritsen L, van Dalen M, Williams A, Cuijpers P, Bockting C. 2020. Prospective biomarkers of major depressive disorder: a systematic review and meta-analysis. Molecular Psychiatry 25:321–338. DOI: 10.1038/s41380-019-0585-z

Kraus C, Kadriu B, Lanzenberger R, Zarate Jr. CA, Kasper S. 2019. Prognosis and improved outcomes in major depression: a review. Translational Psychiatry 9:1–17. DOI: 10.1038/s41398-019-0460-3

Li Y, Ngom A. 2013. The non-negative matrix factorization toolbox for biological data mining. Source Code for Biology and Medicine 8:1–15. DOI: 10.1186/1751-0473-8-10

Liang S, Wang Q, Kong X, Deng W, Yang X, Li Xiaojing, Zhang Z, Zhang J, Zhang C, Li Xin-min, Ma X, Shao J, Greenshaw AJ, Li T. 2019. White Matter Abnormalities in Major Depression Biotypes Identified by Diffusion Tensor Imaging. Neuroscience Bulletin 35:867–876. DOI: 10.1007/s12264-019-00381-w

Liao Y, Huang X, Wu Q, Yang C, Kuang W, Du M, Lui S, Yue Q, Chan RCK, Kemp GJ, Gong Q. 2013. Is depression a disconnection syndrome? Meta-analysis of diffusion tensor imaging studies in patients with MDD. Journal of Psychiatry and Neuroscience 38:49–56. DOI: 10.1503/jpn.110180, PMID: 22691300

Long F, Chen Y, Zhang Q, Li Q, Wang Yaxuan, Wang Yitian, Li H, Zhao Y, McNamara RK, DelBello MP, Sweeney JA, Gong Q, Li F. 2025. Predicting treatment outcomes in major depressive disorder using brain magnetic resonance imaging: a meta-analysis. Molecular Psychiatry 30:825–837. DOI: 10.1038/s41380-024-02710-6

Luttenbacher I, Phillips A, Kazemi R, Hadipour AL, Sanghvi I, Martinez J, Adamson MM. 2022. Transdiagnostic role of glutamate and white matter damage in neuropsychiatric disorders: A Systematic Review. Journal of Psychiatric Research 147:324–348. DOI: 10.1016/j.jpsychires.2021.12.042

Mao Y, Fan L, Feng C, Dai Z. 2025. Predicting responses of neuromodulation and psychotherapies for major depressive disorder: A coordinate-based meta-analysis of functional magnetic resonance imaging studies. Neuroscience & Biobehavioral Reviews 172:106120. DOI: 10.1016/j.neubiorev.2025.106120

Marzola P, Melzer T, Pavesi E, Gil-Mohapel J, Brocardo PS. 2023. Exploring the Role of Neuroplasticity in Development, Aging, and Neurodegeneration. Brain Sciences 13:1610. DOI: 10.3390/brainsci13121610

Menon V. 2011. Large-scale brain networks and psychopathology: a unifying triple network model. Trends in Cognitive Sciences 15:483–506. DOI: 10.1016/j.tics.2011.08.003

Pascalau R, Popa Stănilă R, Sfrângeu S, Szabo B. 2018. Anatomy of the Limbic White Matter Tracts as Revealed by Fiber Dissection and Tractography. World Neurosurgery 113:e672–e689. DOI: 10.1016/j.wneu.2018.02.121

Pasternak O, Kelly S, Sydnor VJ, Shenton ME. 2018. Advances in microstructural diffusion neuroimaging for psychiatric disorders. NeuroImage 182:259–282. DOI: 10.1016/j.neuroimage.2018.04.051

Schaefer A, Kong R, Gordon EM, Laumann TO, Zuo X-N, Holmes AJ, Eickhoff SB, Yeo BTT. 2018. Local-Global Parcellation of the Human Cerebral Cortex from Intrinsic Functional Connectivity MRI. Cerebral Cortex 28:3095–3114. DOI: 10.1093/cercor/bhx179

Schmahmann JD, Guell X, Stoodley CJ, Halko MA. 2019. The Theory and Neuroscience of Cerebellar Cognition. Annual Review of Neuroscience 42:337–364. DOI: 10.1146/annurev-neuro-070918-050258

Smith RE, Tournier J-D, Calamante F, Connelly A. 2015. The effects of SIFT on the reproducibility and biological accuracy of the structural connectome. NeuroImage 104:253–265. DOI: 10.1016/j.neuroimage.2014.10.004

Smith RE, Tournier J-D, Calamante F, Connelly A. 2013. SIFT: Spherical-deconvolution informed filtering of tractograms. NeuroImage 67:298–312. DOI: 10.1016/j.neuroimage.2012.11.049

Smith SM, Jenkinson M, Johansen-Berg H, Rueckert D, Nichols TE, Mackay CE, Watkins KE, Ciccarelli O, Cader MZ, Matthews PM, Behrens TEJ. 2006. Tract-based spatial statistics: Voxelwise analysis of multi-subject diffusion data. NeuroImage 31:1487–1505. DOI: 10.1016/j.neuroimage.2006.02.024

Thase ASWTJAPP van WE van der MHHSME. 2009. Early Improvement in the First 2 Weeks as a Predictor of Treatment Outcome in Patients With Major Depressive Disorder: A Meta-Analysis Including 6562 Patients. The Journal of Clinical Psychiatry 70:5290. DOI: 10.4088/JCP.07m03780

Tian Y, Margulies DS, Breakspear M, Zalesky A. 2020. Topographic organization of the human subcortex unveiled with functional connectivity gradients. Nature Neuroscience 23:1421–1432. DOI: 10.1038/s41593-020-00711-6

Tokuda T, Yoshimoto J, Shimizu Y, Okada G, Takamura M, Okamoto Y, Yamawaki S, Doya K. 2018. Identification of depression subtypes and relevant brain regions using a data-driven approach. Scientific Reports 8:14082. DOI: 10.1038/s41598-018-32521-z

Tournier J-D, Smith R, Raffelt D, Tabbara R, Dhollander T, Pietsch M, Christiaens D, Jeurissen B, Yeh C-H, Connelly A. 2019. MRtrix3: A fast, flexible and open software framework for medical image processing and visualisation. NeuroImage 202:116137. DOI: 10.1016/j.neuroimage.2019.116137

Tustison NJ, Avants BB, Cook PA, Zheng Y, Egan A, Yushkevich PA, Gee JC. 2010. N4ITK: Improved N3 Bias Correction. IEEE Transactions on Medical Imaging 29:1310–1320. DOI: 10.1109/TMI.2010.2046908

Veraart J, Novikov DS, Christiaens D, Ades-aron B, Sijbers J, Fieremans E. 2016. Denoising of diffusion MRI using random matrix theory. NeuroImage 142:394–406. DOI: 10.1016/j.neuroimage.2016.08.016

Whelan R, Garavan H. 2014. When Optimism Hurts: Inflated Predictions in Psychiatric Neuroimaging. Biological Psychiatry 75:746–748. DOI: 10.1016/j.biopsych.2013.05.014

Xia CH, Ma Z, Ciric R, Gu S, Betzel RF, Kaczkurkin AN, Calkins ME, Cook PA, García de la Garza A, Vandekar SN, Cui Z, Moore TM, Roalf DR, Ruparel K, Wolf DH, Davatzikos C, Gur RC, Gur RE, Shinohara RT, Bassett DS, Satterthwaite TD. 2018. Linked dimensions of psychopathology and connectivity in functional brain networks. Nature Communications 9:3003. DOI: 10.1038/s41467-018-05317-y

Yildiz O, Kabatas S, Yilmaz C, Altinors N, Agaoglu B. 2010. Cerebellar mutism syndrome and its relation to cerebellar cognitive and affective function: Review of the literature. Annals of Indian Academy of Neurology 13:23. DOI: 10.4103/0972-2327.61272

Zalesky A, Fornito A, Harding IH, Cocchi L, Yücel M, Pantelis C, Bullmore ET. 2010. Whole-brain anatomical networks: Does the choice of nodes matter? NeuroImage 50:970–983. DOI: 10.1016/j.neuroimage.2009.12.027

Zemmoura I, Burkhardt E, Herbet G. 2021. The inferior longitudinal fasciculus: anatomy, function and surgical considerations - Journal of Neurosurgical Sciences 2021 December;65(6):590-604. Journal of Neurosurgical Sciences 65:590–604. DOI: 10.23736/S0390-5616.21.05391-1

